# Renal Profile of Herbal Medicine Users versus Non-users: an Exploratory Cross-sectional Study in a Family Medicine Clinic in Nigeria

**DOI:** 10.1101/2024.01.30.24301915

**Authors:** Afisulahi Abiodun Maiyegun, Mark Divine Akangoziri, Bukar Alhaji Grema, Yahkub Babatunde Mutalub, Farida Buhari Ibrahim

## Abstract

**Background:** A major risk of herbal medicine is the potential for toxicity and serious side effects. Unlike orthodox medicine, herbs are usually consumed without prior safety assessment via clinical studies. This means the clinical effects of herbs may be detected only through an assessment of health parameters of consumers.

**Objective:** To evaluate participants’ renal profile, comparing that of herb users with non-users, thereby identifying any differences between the two groups.

**Design:** Cross-sectional study.

**Setting:** Primary care clinic of a teaching hospital in Nigeria

**Participants:** 341 patients participated in the study. Of these, 60% were female and 80 % were of Hausa/Fulani ethnicity. All adult patients attending the clinic were eligible. Very sick patients were excluded from the research.

**Primary outcome:** estimated glomerular filtration rate, microalbuminuria and proteinuria among patients who reported using herbs and those who did not report any use.

**Secondary outcome:** electrolyte, urea, and creatinine levels among study participants.

**Results:** The mean eGFR for participants who used herbs was 87.47 ± 25.44 ml/min/1.73m^2^ and 84.76 ± 25.49 ml/min/1.73m^2^ for those who never used herbs (P value 0.48). Proteinuria ≥0.3g/l was present in 29(8.50%) of participants, and microalbuminuria ≥30mg/l in 138 (40.47%). There was no statistically significant difference in the renal parameters of both groups.

**Conclusion:** The study could not establish harmful effect of the herbs reported on kidney function. Further studies in human subjects of the effect of herbal medicine on the kidney and other systems are suggested.

**Article Summary:** *Strengths and Limitations:*

- This study provides empiric evidence to compare renal function among users of herbal medicine and others.
- The study provides a detailed evaluation of renal function in the participants using estimated glomerular filtration rate, as well as urine protein and microalbumin estimation.
- It identifies the common herbs used in a primary care clinic in the most populous black nation.
- A major limitation of the study is that the quantity and frequency of herbal medicine use were not measured.

## INTRODUCTION

Herbs, like orthodox medicine, have potential systemic side effects and toxicities. However, evidence for herbal toxicity in clinical scenarios are likely to be based mainly on case reports and patient anecdotes, because, unlike conventional drugs, herbs do not typically undergo clinical trials before their use.[1,2] Nevertheless, many studies support the use of herbal medicines even for severe cases like liver disease, diabetes and hypertension, suggesting the therapeutic effects of herbs could sometimes outweigh the toxicities.[3–5]

These divergent perspectives highlight the inconclusive nature of the argument on herbal medicine use, and demands more rigorous research to determine the benefits and harms herbal medicine use entails.[6] As the use of herbs for treatment continues to thrive both in the developed and the developing world, research about ongoing herbal medicine use in the clinics and community are, therefore, necessary. Importantly, such ‘surveillance’ research could help avoid or aid early detection of herb-related complications such as Balkan nephropathy-where herbs in the long history of certain communities were found to contain aristolochic acid associated with nephropathy and urothelial cancer – and Chinese herbal nephropathy.[7,8]

Assessment of clinical and biochemical parameters of patients based on herbal medicine use has rarely been reported in Nigeria. Because of the importance of the kidneys in the metabolic and cardiovascular wellbeing, and the enormous socioeconomic burden associated with renal morbidity and mortality, this study aimed to assess the renal profile of patients attending a general outpatient clinic to determine any association with herbal medicine use.

## METHODS

The study was conducted in the general outpatient unit of Abubakar Tafawa Balewa University Teaching Hospital (ATBUTH), Bauchi. It took place between September and December 2018. The study population were adult patients (at least 18 years old) attending the general outpatient clinic. All patients attending the general outpatient clinic during the study period were eligible for the study. Patients who were very sick or declined consent were excluded from the study. Sample size for the study was determined using the Cochran’s formula: N = Z^2^pq/d^2^, where N = minimum sample size, Z = confidence level at 95% (1.96), p = 0.67, being the prevalence of herbal medicine use from a previous study in Nigeria (Oreagba et al), q = (1-p), and d = level of precision (0.05).[9] The sampling size was 341. Prospective participants were selected through systematic random sampling, and their consent sought and obtained. About 3,000 adults attend the General Outpatient Clinic monthly (excluding Saturdays and Sundays), which gave a sampling frame of 9,000 over the three-month period of the study (3,000 ×3). All patients, who were selected by random sampling, agreed to participate in the study.

In the first part of the research, an interviewer-administered, semi-structured questionnaire was used to collect information on participants’ biodata, medical history, and use of herbal medicine. In this second part, urine and venous blood samples were collected from study participants. The urine was tested for protein and microalbuminuria, using the DIRUI H-13Cr^®^ urine strip manufactured by the Dirui Industrial Company Limited, Turkey. The blood sample was analysed for electrolytes, urea and creatinine, and estimated glomerular filtration rate (eGFR) values were calculated using the Chronic Kidney Disease Epidemiology Collaboration (CKD-EPI) equation:[10] GFR = 141 ×min (Scr/κ, 1)^α^ × max(Scr/κ, 1)^-1.209^ × 0.993^Age^ × 1.018[if female] × 1.159[if black]

Where Scr = standardised serum creatinine in mg/dl

κ = 0.7 for females and 0.9 for males

α = −0.329 for females and −0.411 for males

min indicates the minimum of Scr/κ or 1

max indicates the maximum of Scr/κ or 1

Albuminuria was present if urine albumin was ≥ 30mg/l and proteinuria present if urine protein ≥ 0.3g/l. These values were compared between participants who used herbal medicine and those who did not. To control for confounding, subgroup analysis was done for participants with diabetes and hypertension, as these two groups had disease-related risks for poor renal outcome.

Epi Info version 7.2 was used for the collation, storage and analysis of the study data. Continuous variables like the estimated glomerular filtration rate were summarised using the mean and standard deviation, while categorical variables like the results of urinalysis were summarised using percentages. In comparing the study outcomes between participants who used herbs and those who did not, the Student t-test was used for continuous variables like eGFR, and Chi square was used for categorical variables like urinalysis result.

Ethical approval for the study (Assigned Number: 03/03/2018) was granted by the Research and Ethics Committee of Abubakar Tafawa Balewa University, Bauchi, where the study was carried out.

## RESULTS

Sociodemographic characteristics of the participants are summarised in Table 1. The modal age group was 25 - 44 years.

**Table 1.** Sociodemographic Characteristics of the Respondents.

| Sociodemographic variable | Frequency | Percentage (%) |
| --- | --- | --- |
| **Age** |  |  |
| 18-24 | 56 | 16.42 |
| 25-44 | 168 | 49.27 |
| 45-64 | 98 | 28.74 |
| 65-85 | 19 | 05.57 |
| **Gender** |  |  |
| Male | 137 | 40.18 |
| Female | 204 | 59.82 |
| **Tribe** |  |  |
| Hausa/Fulani | 272 | 79.77 |
| Igbo | 8 | 02.34 |
| Yoruba | 6 | 01.76 |
| Others | 55 | 16.13 |
| **Religion** |  |  |
| Christianity | 48 | 14.08 |
| Islam | 293 | 85.92 |
| **Education** |  |  |
| No formal education | 75 | 22.00 |
| Primary education | 47 | 13.78 |
| Secondary education and above | 219 | 64.22 |
| **Occupation** |  |  |
| Public Servant | 102 | 29.92 |
| Self-employed | 118 | 34.60 |
| Unemployed | 121 | 35.48 |
| **Marital Status** |  |  |
| Currently Married | 258 | 75.66 |
| Not currently Married | 83 | 24.34 |
| **Average family income (Naira)** |  |  |
| <50,000 | 122 | 35.78 |
| >50,000-<100,000 | 51 | 14.96 |
| ≥100,000 | 55 | 16.13 |
| Undeclared | 113 | 33.13 |
| \$1 equivalent to 324.2 Naira in 2018 | | |

Of the total participants, 118 (34.60%) had hypertension and 26 (7.62%) had diabetes. In the study, 291 participants had used herbal medicine before, resulting in a lifetime prevalence of 85.34%. Of these, 14 (4.81%) had used it throughout the preceding twelve months. Table 2 shows the different herbs reported by the study participants.

**Table 2.**
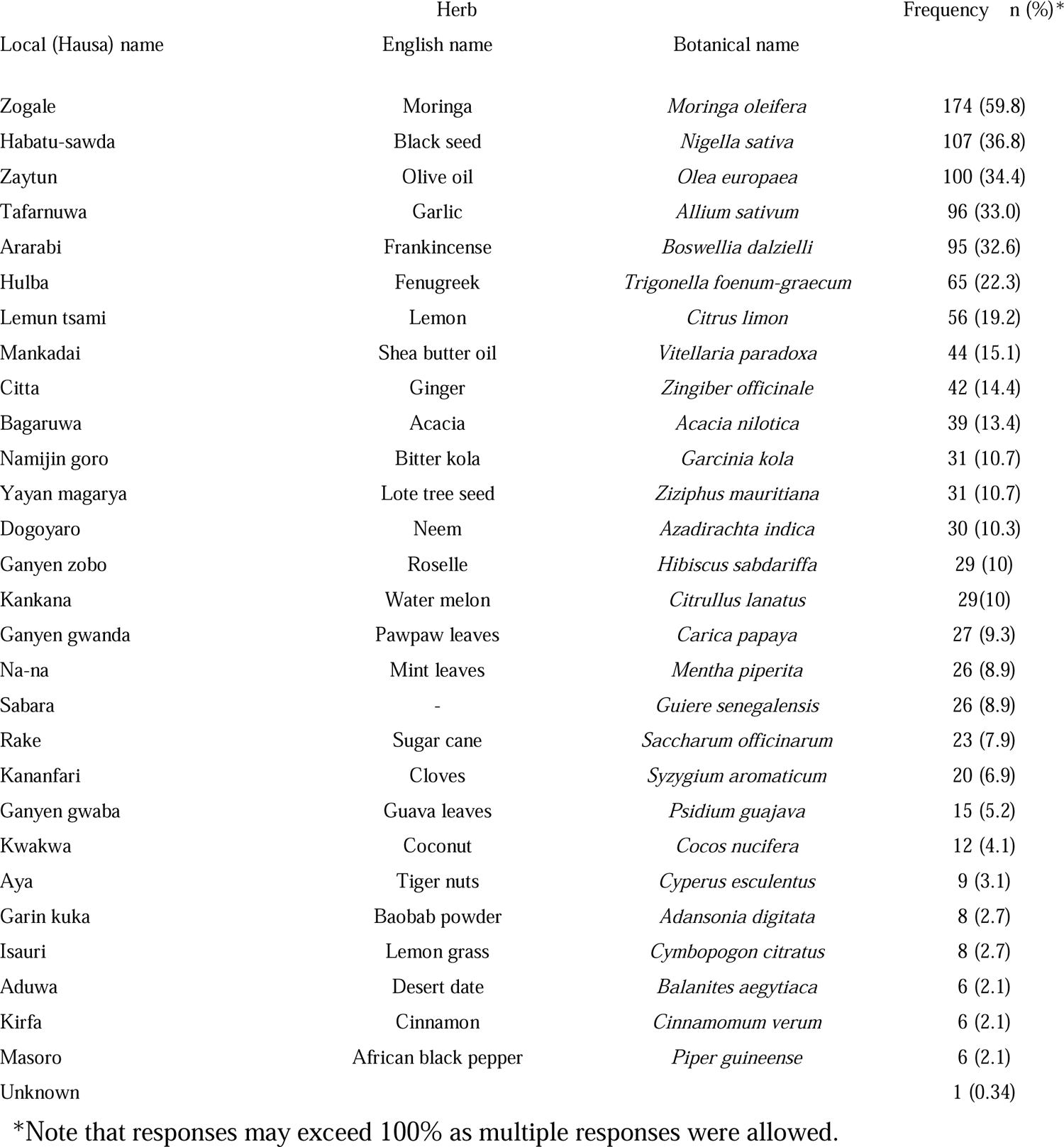
Herbs Used by Participant in the Study.

| Local (Hausa) name | Herb |  | Frequency n (%)* |
| --- | --- | --- | --- |
|  | English name | Botanical name |  |
| Zogale | Moringa | *Moringa oleifera* | 174 (59.8) |
| Habatu-sawda | Black seed | *Nigella sativa* | 107 (36.8) |
| Zaytun | Olive oil | *Olea europaea* | 100 (34.4) |
| Tafarnuwa | Garlic | *Allium sativum* | 96 (33.0) |
| Ararabi | Frankincense | *Boswellia dalzielli* | 95 (32.6) |
| Hulba | Fenugreek | *Trigonella foenum-graecum* | 65 (22.3) |
| Lemun tsami | Lemon | *Citrus limon* | 56 (19.2) |
| Mankadai | Shea butter oil | *Vitellaria paradoxa* | 44 (15.1) |
| Citta | Ginger | *Zingiber officinale* | 42 (14.4) |
| Bagaruwa | Acacia | *Acacia nilotica* | 39 (13.4) |
| Namijin goro | Bitter kola | *Garcinia kola* | 31 (10.7) |
| Yayan magarya | Lote tree seed | *Ziziphus mauritiana* | 31 (10.7) |
| Dogoyaro | Neem | *Azadirachta indica* | 30 (10.3) |
| Ganyen zobu | Roselle | *Hibiscus sabdariffa* | 29 (10) |
| Kankana | Water melon | *Citrullus lanatus* | 29 (10) |
| Ganyen gwanda | Pawpaw leaves | *Carica papaya* | 27 (9.3) |
| Na-na | Mint leaves | *Mentha piperita* | 26 (8.9) |
| Sabara | - | *Guiera senegalensis* | 26 (8.9) |
| Rake | Sugar cane | *Saccharum officinarum* | 23 (7.9) |
| Kananfari | Cloves | *Syzygium aromaticum* | 20 (6.9) |
| Ganyen gwaba | Guava leaves | *Psidium guajava* | 15 (5.2) |
| Kwakwa | Coconut | *Cocos nucifera* | 12 (4.1) |
| Aya | Tiger nuts | *Cyperus esculentus* | 9 (3.1) |
| Garin kuka | Baobab powder | *Adansonia digitata* | 8 (2.7) |
| Isauri | Lemon grass | *Cymbopogon citratus* | 8 (2.7) |
| Aduwa | Desert date | *Balanites aegytiaca* | 6 (2.1) |
| Kirfa | Cinnamon | *Cinnamomum verum* | 6 (2.1) |
| Masoro | African black pepper | *Piper guineense* | 6 (2.1) |
| Unknown |  |  | 1 (0.34) |

\*Note that responses may exceed 100% as multiple responses were allowed.

### Microalbuminuria among Study Participants

Among all the participants in the study, 203(59.53%) had microalbuminuria level less than 30mg/l and 138 (40.47%) had microalbuminuria ≥30mg/l. Among participants with hypertension, 63(53.39%) had microalbuminuria <30mg/l and 55(46.61%) had microalbuminuria ≥30mg/l. Among participants with diabetes, 12(46.15%) had microalbuminuria <30mg/l and 14(53.85%) had microalbuminuria ≥30 mg/l. Microalbuminuria was not associated with herbal medicine use (Supplementary Table 1)

### Proteinuria among Study Participants

Among all the participants in the study, 312(91.50%) had proteinuria level less than 0.3g/l and only 29(8.50%) had proteinuria ≥0.3g/l. Among participants with hypertension, 103(87.29%) had proteinuria <0.3g/l and 15(12.71%) had proteinuria ≥0.3g/l. Among participants with diabetes, 23(88.46%) had proteinuria <0.3g/l and 3(11.54%) had proteinuria ≥0.3 mg/l. Proteinuria was not associated with herbal medicine use (Supplementary Table 2).

### Serum Electrolyte and Estimated Glomerular Filtration Rate (eGFR) among Study Participants

Tables 3-5 show the results of the renal function test for the participants. There was no statistically significant difference in the parameters of the renal function test between herbal medicine users and non-users. Overall (n=341), forty-one participants (12.02%) had eGFR less than 60ml/minute/1.73m^2^. Among participants with hypertension (n=118), twenty-eight participants (23.73%) had eGFR less than 60ml/minute/1.73m^2^, and among participants with diabetes (n=26), eleven participants (42.31%) had eGFR less than 60ml/minute/1.73m^2^.

**Table 3.** Serum Electrolyte and Estimated Glomerular Filtration Rate (eGFR) among All Study.

| Electrolyte<br>(mmol/l) | Total(n=341) | Used Herbs(n=291) | No Herbs (n=50) | Mean<br>Difference | Confidence<br>Interval |
| --- | --- | --- | --- | --- | --- |
|  | Mean±S. D | Mean±S. D | Mean±S. D |  |  |
| Sodium | 138.33±4.13 | 138.28±4.04 | 138.60±4.61 | -0.32 | -1.57, 0.18 |
| Potassium | 3.96±1.86 | 3.87±0.46 | 3.82±0.37 | 0.05 | -0.09, 0.18 |
| Chloride | 97.81±4.54 | 97.76±4.43 | 98.14±5.20 | -0.38 | -1.75, 0.99 |
| Bicarbonate | 26.17±2.63 | 26.15±2.68 | 26.24±2.31 | -0.09 | -0.88, 0.71 |
| Urea | 4.26±5.16 | 4.32±5.50 | 3.91±2.23 | 0.41 | -1.15, 1.96 |
| Creatinine* | 102.65±61.33 | 102.27±60.60 | 104.86±66.02 | -2.59 | -21.09,<br>15.90 |
| eGFR<sup>+</sup> | 87.07±25.43 | 87.47±25.44 | 84.76±25.49 | 2.71 | -5.08,<br>10.50 |
| eGFR<sup>+</sup> | Assessment for CKD |  |  | X<sup>2</sup> | p value |
| ≥60 |  | 256(85.33%) | 44(14.67%) | 0.00 | 1.00 |
| <60 |  | 35(85.37%) | 6(14.63%) |  |  |

\* μmol/l
+mol/min/1.73m<sup>2</sup>
Df=339
SD (Standard Deviation)

**Table 4.** Serum Electrolyte and Estimated Glomerular Filtration Rate (eGFR) among Study Participants with Hypertension.

| Electrolyte<br>(mmol/l) | Total (n=118) | Used Herbs(n=98) | No Herbs (n=20) | Mean<br>Difference | Confidence<br>Interval |
| --- | --- | --- | --- | --- | --- |
|  | Mean ± S.D | Mean ± S.D | Mean±S.D |  |  |
| Sodium | 138.52±4.04 | 138.62±3.68 | 138.00±5.59 | 0.62 | -1.35, 2.59 |
| Potassium | 3.80±0.52 | 3.81±0.55 | 3.76±0.41 | 0.05 | -0.20, 0.31 |
| Chloride | 97.56±4.47 | 97.80±4.23 | 96.40±5.51 | 1.40 | -0.77, 3.56 |
| Bicarbonate | 26.25±2.71 | 26.19±2.79 | 26.55±2.39 | -0.36 | -1.68, 0.97 |
| Urea | 4.76±4.20 | 4.79±4.39 | 4.65±3.18 | 0.14 | -1.91, 2.19 |
| Creatinine* | 116.96±96.53 | 114.83±96.17 | 127.40±100.15 | -12.57 | -59.63,<br>34.48 |
| eGFR<sup>+</sup> | 74.75±24.42 | 75.93±25.04 | 68.95±20.64 | 6.98 | -4.87,<br>18.83 |
| eGFR<sup>+</sup> | Assessment for CKD |  |  | X<sup>2</sup> | p value |
| >60 |  | 75(83.33%) | 15(16.67%) | 0.02 | 0.88 |
| <60 |  | 23(82.14%) | 5(17.86%) |  |  |
| * μmol/l |  |  |  |  |  |
| +mol/min/1.73m<sup>2</sup> |  |  |  |  |  |
| Df=116 |  |  |  |  |  |
| SD (Standard Deviation) |  |  |  |  |  |

**Table 5.** Serum Electrolyte and Estimated Glomerular Filtration Rate (eGFR) among Study Participants with Diabetes.

| Electrolyte (mmol/l) | Total (n=26) | Used Herbs(n=22) | No Herbs (n=4) | Mean Difference | Confidence Interval |
| --- | --- | --- | --- | --- | --- |
|  | Mean±S.D | Mean±S.D | Mean±S.D |  |  |
| Sodium | 137.85±4.24 | 137.18±4.26 | 141.50±1.29 | -4.32 | -8.82, 0.18 |
| Potassium | 4.00±0.47 | 4.03±0.48 | 3.83±0.39 | 0.21 | -0.32, 0.74 |
| Chloride | 96.04±5.24 | 95.41±5.42 | 99.50±2.08 | -4.09 | -9.8, 1.66 |
| Bicarbonate | 25.92±2.94 | 25.81±3.13 | 26.50±1.73 | -0.68 | -4.03, 2.67 |
| Urea | 4.63±2.60 | 4.90±2.73 | 3.10±0.87 | 1.80 | -1.08, 4.69 |
| Creatinine* | 109.04±33.99 | 110.05±36.39 | 103.50±17.52 | 6.55 | -32.27, 45.36 |
| eGFR<sup>+</sup> | 69.15±21.41 | 69.50±22.08 | 67.25±19.97 | 2.25 | -22.24, 26.74 |
| eGFR<sup>+</sup> |  | Assessment for CKD |  | X<sup>2</sup> | F value |
| >60 |  | 13(86.67%) | 2(13.33%) | 0.11 | 1.00 |
| <60 |  | 9(81.82%) | 2(18.18%) |  |  |
| * μmol/l<br>+mol/min/1.73m<sup>2</sup><br>Df=24<br>SD (Standard Deviation) |  |  |  |  |  |

## DISCUSSION

This was a study investigating the renal function of 341 patients with respect to herbal medicine use in a general outpatient clinic of a tertiary hospital in Nigeria. The study showed that 40.47% of the study population had significant microalbuminuria, with expectedly higher values among those with hypertension and diabetes (Supplementary Table 1). In a family practice in Nigeria, Afolabi et al. found a similar 44.4% microalbuminuria rate among outpatients.[11] As microalbuminuria is an independent indicator and earlier marker of renal damage, this high level suggests significant latent renal damage among the study population, and mandates a more intensive approach to screening for and managing renal-related conditions, including hypertension and diabetes.

In contrast to the high level of microalbuminuria observed in the study, only 8.50% of the study population had proteinuria, confirming that microalbuminuria usually heralds proteinuria, the more obvious marker of renal damage. The rate of proteinuria in this study is higher than that reported by Ulasi et al. in South-East Nigeria (3.7%).[12] This could be explained by the fact that the latter study had lower prevalence of hypertension (26.1% vs 34.60%) and diabetes (5.90%vs 7.62%), two known causes of proteinuria.

Although the means of the serum electrolyte, urea and creatinine for all the study participants were within normal range (Tables 3-5), the prevalence of chronic kidney disease (eGFR<60ml/min/1.73m^2^) in the study was 12.02%, similar to that in other community and practice-based studies in Nigeria (11.4% to 26%), China (10.8%) and Taiwan (11.9%), though slightly below the prevalence reported in the Middle East (5.2% to 10.6%) [11,13–16] This finding emphasises the high prevalence of CKD in the Nigerian population, and the need to focus on renal health, especially with the considerable evidence of early renal impairment as evidenced by the significant microalbuminuria (40.47%), which suggests that a high proportion of the population with presumed normal renal function are only in the latent stage of the disease, and could, without appropriate medical intervention, progress to frank chronic kidney disease in the future.

The study failed to show any relationship between herbal medicine use and renal status among the study participants. The reduction in eGFR observed in the hypertension and diabetes is not unexpected, since these two conditions have marked renal impact. However, this, too, was not found to be statistically associated with herbal medicine use by the participants. The failure to establish a significant difference between the renal profile of those who used herbs and those who did not in this study (using eGFR, proteinuria and microalbuminuria) may be due to the safety profile of many of the common herbs, such as moringa, *Nigella sativa*, olive oil, ginger, lemon, pumpkin, sugar cane, coconut, tiger nuts, guava and shea butter oil, all of which are known edible substances.

### Limitations

The quantity and frequency of herbal medicine use was not quantified in this study since numerous herbs had been used by the participants over different periods. This would be better addressed in a clinical trial, which may not be ethically permissible as herbs are not the standard of care in Nigeria.

## CONCLUSIONS

This study identified a high prevalence of chronic kidney disease (12.02%) and early renal impairment (significant microalbuminuria of 40.47%) in the study population. However, it did not find any statistically significant association between use of herbal medicines and renal impairment. It suggests that the effect of herbs commonly used by patients attending general outpatient clinics in Northern Nigeria may be relatively safe to the kidneys. This may not be generalisable to other parts of the country, or to other countries, because the kinds of herbs used in those settings are likely to differ from that reported in this study. However, the study underlines the need for identifying the herbs in common use among the patient population to prevent unsubstantiated assumptions of herbal medicine harm or benefits. Finally, it demonstrates the potential ongoing renal damage in the patient population. We encourage similar studies in other parts of the world to understand the effect of patients’ herbal choices on their health.

## Data Availability

All data produced in the present study are available upon reasonable request to the authors.

https://doi.org/10.6084/m9.figshare.25062332

## Acknowledgements

Late Dr AbdulRazak Toyin, former Coordinator of Laboratory Services, ATBUTH, Bauchi, for providing invaluable assistance with laboratory logistics during the research. Mr Ibrahim A. Ibrahim, formerly of the general outpatient unit of the Family Medicine Department, for providing oversight of the workplace during the research.

## Author Statement

Dr. Afisulahi Abiodun Maiyegun: Conceptualization of the Study, Data Collection, Analysis, Interpretation, Drafting of the Manuscript, and Final Editing. Dr. Mark Divine Akangoziri: Conceptualization of the Study, Interpretation, Manuscript Review Dr. Bukar Alhaji Grema: Conceptualization of the Study, Interpretation, Manuscript Review, Dr. Yahkub Babatunde Mutalub: Conceptualization of the Study, Interpretation, Manuscript Review, Farida Buhari Ibrahim: Conceptualization of the Study, Data Collection, Manuscript Review All authors approve the final manuscript. The dataset is available in Fighshare repository. DOI: 10.6084/m9.figshare.25062332

## REFERENCES

1. Özkan G, Ulusoy Ş. A case of renal failure developing in association with African mango consumption [Internet]. Int J Clin Exp Med. 2015; 8(4): 6374–6378 Available from: www.ijcem.com/

2. Akpan EE, Ekrikpo UE. Acute renal failure induced by Chinese Herbal Medication in Nigeria. 2015;2015: 150204.

3. Saleem TSM, Chetty CM, Ramkanth S, et al. Hepatoprotective Herbs – A Review. Int J Res Pharm Sci. 2010;1(1):1–5.

4. Tabassum N, Ahmad F. Role of natural herbs in the treatment of hypertension. Pharmacogn Rev. 2011;5(9):30–40.

5. Osadebe P, Odoh E, Uzor P. Natural products as potential sources of antidiabetic drugs. Br J Pharm Res. 2014;4(17):2075–95.

6. Pelkonen O, Xu Q, Fan TP. Why is research on herbal medicinal products important and how can we improve its quality? J Tradit Complement Med. 2014;4(1):1–7.

7. Grollman AP. Aristolochic acid nephropathy: Harbinger of a global iatrogenic disease. Environ Mol Mutagen. 2013; 54:1–7.

8. Yang B, Xie Y, Guo M, et al. Nephrotoxicity and Chinese herbal medicine. Clin J Am Soc Nephrol. 2018; 13(10): 1605–11.

9. Oreagba IA, Oshikoya KA, Amachree M. Herbal medicine use among urban residents in Lagos, Nigeria. BMC Complement Altern Med. 2011;11(1):117.

10. Levey AS, Stevens LA, Schmid CH, et al. A new equation to estimate glomerular filtration rate. Ann Intern Med. 2009;150(9):604–12.

11. Afolabi MO, Abioye-Kuteyi EA, Arogundade FA, et al. Prevalence of chronic kidney disease in a Nigerian family practice. South African Family Practice. 2014;51(2):132–7.

12. Ulasi II, Ijoma CK, Onodugo OD, et al. Towards prevention of chronic kidney disease in Nigeria: a community-based study in Southeast Nigeria. Kidney Int Suppl (2011). 2013; 3:195–201.

13. Zhang L, Wang F, Wang L, et al. Prevalence of chronic kidney disease in China: a cross-sectional survey. The Lancet. 2010;379(9818):815–22.

14. Hwang S-J, Tsai J-C, Chen H-C. Epidemiology, impact and preventive care of chronic kidney disease in Taiwan. Nephrology. 2010; 15:3–9.

15. Chukwuonye II, Ogah OS, Anyabolu EN, et al. Prevalence of chronic kidney disease in Nigeria: systematic review of population-based studies. Int J Nephrol Renovasc Dis. 2018; 11:165–72.

16. Amouzegar A, Abu-Alfa AK, Alrukhaimi MN, et al. International Society of Nephrology Global Kidney Health Atlas: structures, organization, and services for the management of kidney failure in the Middle East. Kidney Int Suppl. 2021; 11(2): e47–e56.

