## Supplemental Tables for "Renal Profile of Herbal Medicine Users versus Non-users: an Exploratory Cross-sectional Study in a Family Medicine Clinic in Nigeria"

SUPPLEMENTARY TABLES

Supplementary Table 1 Microalbuminuria among Study Participants

| Herbal Use | Microalbuminuria (mg/l) | | Χ^2^ | p value |
| --- | --- | --- | --- | --- |
|  | <30 | ≥30 |  |  |
|  | All study participants (n=341) | |  |  |
| Yes | 169 (58.08%) | 122 (41.92%) | 1.74 | 0.19 |
| No | 34 (68.00%) | 16 (32.00%) |  |  |
|  | Participants with hypertension(n=118) | | |  |
| Yes | 50 (51.02%) | 48 (48.98%) | 1.30 | 0.25 |
| No | 13 (65.00%) | 7 (35.00%) |  |  |
|  | Participants with diabetes mellitus (n=26) | | |  |
| Yes | 10 (45.45%) | 12 (54.55%) |  | 0.87* |
| No | 2 (50.00%) | 2 (50.00%) |  |  |

* Fisher’s exact test was used for cells with value <5.

Supplementary Table 2 Proteinuria among Study Participants

| Herbal medicine use | Protein | | Χ^2^ | p value |
| --- | --- | --- | --- | --- |
|  | Negative | Positive |  |  |
|  | All Study Participants (n=341) | |  |  |
| Yes | 267(91.75%) | 24 (8.25%) | 0.17 | 0.68 |
| No | 45 (90.00%) | 5 (10.00%) |  |  |
|  | Participants with hypertension(n=118) | | |  |
| Yes | 85 (86.73%) | 13 (13.27%) | 0.16 | 1.00* |
| No | 18 (90.00%) | 2 (10.00%) |  |  |
|  | Participants with diabetes mellitus (n=26) | | |  |
| Yes | 19 (86.36%) | 3 (13.64%) | 0.62 | 1.00 |
| No | 4 (100.00%) | 0 (0.00%) |  |  |

* Fisher’s exact test was used for cells with value <5.
